# Are remote mental healthcare interventions cost-effective? A systematic review of economic evaluations of remote mental healthcare

**DOI:** 10.1101/2022.12.01.22282817

**Authors:** Amy Clark, Rebecca Appleton, Erika Kalocsanyiova, Evdoxia Gkaintatzi, Paul McCrone

## Abstract

**Background:** Remote interventions known as telemental health care increased in use due to the COVID-19 pandemic when social distancing requirements were in place. Whilst there is some evidence regarding the cost-effectiveness of telemental health prior to the pandemic, there is a need for further evaluation due to the increase in remote care.

**Aims:** To systematically review the literature to explore whether remote mental health care interventions are cost-effective in terms of incremental cost per quality adjusted life year and in relation to condition specific outcomes compared to usual care or an alternative intervention.

**Method:** A multilayer search strategy was conducted to build on the searches of a previous systematic review, as well as including grey literature and economic models. Six databases (PubMed, EMBASE, Cochrane Central, PsychINFO, CINAHL, and EconLit) were searched for literature relating to the cost effectiveness of telemental health. Quality appraisal was conducted for all included studies, and findings were synthesised using narrative synthesis.

**Results:** 7386 studies were identified of which 59 met our inclusion criteria and were included in the synthesis of findings. 45 studies were rated as very good or excellent quality. Of the 59 included studies, 40 indicated that the telemental health intervention was cost-effective, whilst a further 16 suggested the intervention had potential to be cost-effective, but there was some uncertainty in the findings. Three studies reported that the intervention was not cost-effective.

**Conclusions:** This evidence will be used to inform practice in the UK as we respond to and recover from the COVID-19 pandemic.

## Introduction

Remote interventions in mental health care, or “tele-mental health” have been implemented for many years and evaluations often include assessment of cost-effectiveness. Telemental health care is defined as any interventions or modes of working where remote technology (e.g. telephone/video call/instant messaging) is used to facilitate direct communication between staff and service users, between service users and peers, or between mental health professionals in different locations.

Telemental health has the potential to result in benefits for both service users and clinicians. For example, service users have identified increased flexibility as a key advantage of remote care (1), and a recent rapid realist review (2) found that telemental health care can improve access to mental health support for people who may struggle to travel to face-to-face appointments due to disability, anxiety about travel, or caring or work responsibilities.

Reported benefits for staff included an opportunity for more flexible working, less time spent travelling, and communicating with other clinicians at different sites (2, 3). However, it is important to note that telemental health is not suitable for everyone; not all service users may wish for care to be remote and inequalities and digital exclusion should be taken into consideration during the decision to offer mental health care remotely (1, 2). Clinicians have also reported finding telemental health less suitable than face-to-face care when treating trauma, or for service users in crisis or those with psychotic symptoms or severe anxiety (3).

The uptake of telemental health care increased rapidly during the COVID-19 pandemic as services moved from largely face-to-face models of care due to social distancing requirements (4, 5). However, a systematic review conducted during the early stages of the pandemic identified a lack of evidence on the cost-effectiveness of telemental health interventions (4).

Pre-pandemic evidence regarding the cost-effectiveness of telemental health was also inconclusive. An umbrella review found that in studies conducted prior to the COVID-19 pandemic, evidence regarding the cost-effectiveness of telemental health was mixed (6).

Some studies identified lower costs of telemental health care due to savings on travel time for service users, or not needing to take time off work, whereas higher costs in others were attributed to the expense of videoconferencing equipment (6). A further systematic review by Naslund et al (7) identified 26 economic evaluations of telepsychiatry programmes, also prior to the COVID-19 pandemic. They found that 60% reported telepsychiatry programmes to be less expensive and 32% reported they were more expensive than usual care, again primarily due to costs of technology and equipment. This review was limited to peer-reviewed studies that used primary data collection and therefore economic models were excluded.

This proposed review therefore aims to build on Naslund et al (7) and bring together further evidence on economic evaluations of remote mental healthcare. We originally set out to answer the following secondary review questions: (i) Does cost-effectiveness of remote mental health care interventions differ by subgroup? (ii) Does cost-effectiveness of remote mental health care interventions differ by model type? (iii) What is the impact of remote technology for staff communication on costs for the health and social care system? (iv) What is the impact of remote technology for mental healthcare interventions on costs for the health and social care system? (v) How is the amount of contact time between service users and mental health care professionals affected by remote interventions? However, given the data reported in the selected studies only research questions ii and iv will be addressed in this paper.

## Methods

The review was registered on International prospective register of Systematic Reviews (PROSPERO) ID: CRD42020216755. We developed a multilayer search strategy to capture both new evidence published since the completion of the Naslund et al. (7) searches on 16 March 2018 and to cover additional sources of information excluded by these authors, namely grey literature and simulation models. We used the same search criteria as outlined in Naslund et al. (7) to identify any relevant peer reviewed literature published since 16 March 2018. The following six databases were searched on 8-10 December 2020: PubMed (MEDLINE), EMBASE, Cochrane Central, PsychINFO (EBSCOhost), CINAHL, and EconLit^1^.

An example of the search terms used, as per Naslund et al. (7), is reported in Table S1^2^ of the Supplementary Material. To identify economic models, which were excluded by Naslund et al. (7), the same strategy was used but with a different date range (January 2000 – October 2020) and two additional search terms added to Table S1: “model OR simulation”. Grey literature databases were searched from 1 January 2000 to 31 October 2020 for terms related to “tele” “mental health” and “costs” as outlined in Table S2. MedNAR and Google were also searched using the following terms to improve relevance of results: “cost effective” AND (tele* OR telehealth OR telepsychiatry OR remote OR technology OR digital) AND mental. Records identified in the search were uploaded to Rayyan QCRI, a web-based reference manager system for collaborative systematic review, for de-duplication and blinded screening and study selection.

### Study selection criteria

After de-duplication, the results were pre-screened by three student research assistants (MT, SS, and JP). A 25 per cent sample was selected at random and checked for accuracy by the second author (EK). The titles and abstracts of the remaining studies were screened in duplicate by three reviewers (AC, EK and JB) against the eligibility criteria shown in Table 1. Disagreements and records marked as undecided were resolved by discussion or a consensus involving a third author.

**Table 1:**
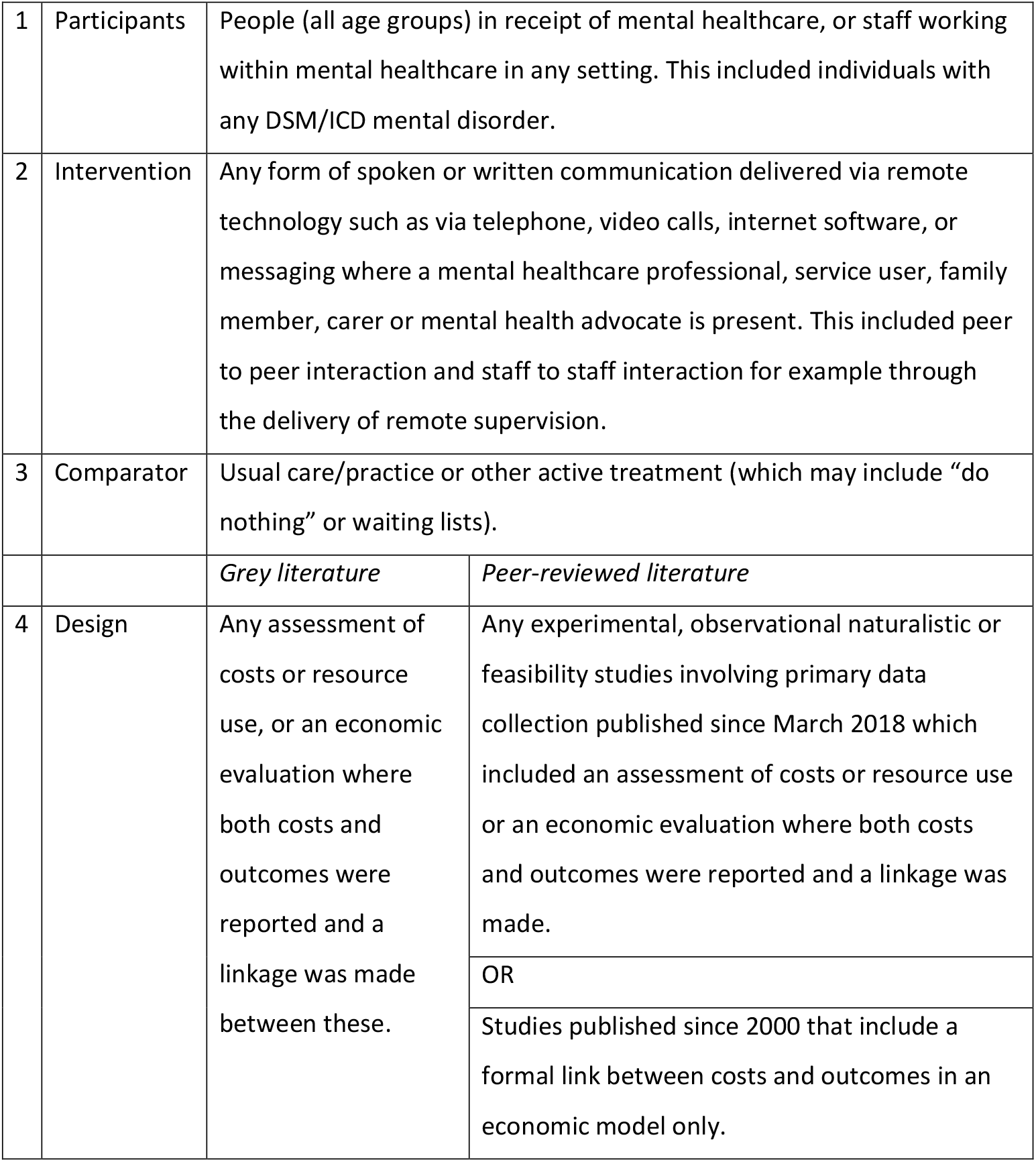
Eligibility criteria.

|  |  |  |  |
| --- | --- | --- | --- |
| 1 | Participants | People (all age groups) in receipt of mental healthcare, or staff working within mental healthcare in any setting. This included individuals with any DSM/ICD mental disorder. |  |
| 2 | Intervention | Any form of spoken or written communication delivered via remote technology such as via telephone, video calls, internet software, or messaging where a mental healthcare professional, service user, family member, carer or mental health advocate is present. This included peer to peer interaction and staff to staff interaction for example through the delivery of remote supervision. |  |
| 3 | Comparator | Usual care/practice or other active treatment (which may include “do nothing” or waiting lists). |  |
|  |  | *Grey literature* | *Peer-reviewed literature* |
| 4 | Design | Any assessment of costs or resource use, or an economic evaluation where both costs and outcomes were reported and a linkage was made between these. | Any experimental, observational naturalistic or feasibility studies involving primary data collection published since March 2018 which included an assessment of costs or resource use or an economic evaluation where both costs and outcomes were reported and a linkage was made. |
|  |  |  | OR |
|  |  |  | Studies published since 2000 that include a formal link between costs and outcomes in an economic model only. |

In the process of abstract and title screening, we identified 21 peer-reviewed cost effectiveness studies published prior to March 2018 which appeared eligible according to the study selection criteria outlined in Naslund et al. (7) but were not included in their review. Given that these studies were identified in grey literature databases not considered by Naslund et al. (7), we decided to assess them for inclusion along with the other studies retained for full text screening.

Where full text was not available, the corresponding authors were contacted. If an answer was not obtained following a reminder and a 3-month waiting period, the study was excluded. Reference list searching was undertaken by the second author (EK). The search and study selection process has been documented in Figure 1 using the Preferred Reporting Items for Systematic Reviews and Meta-Analyses PRISMA guidelines (8).

**Figure 1:**
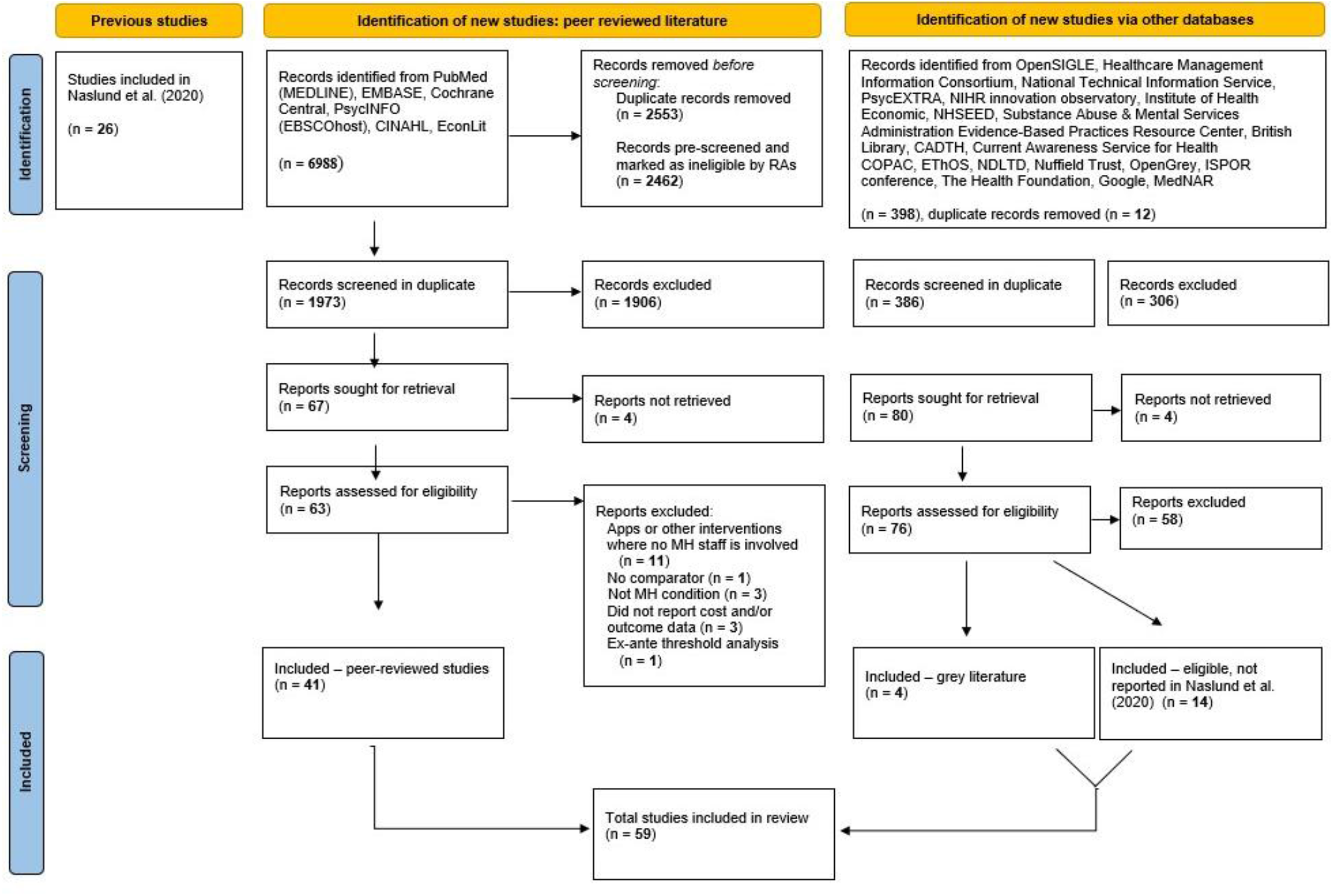
PRISMA flow chart.

### Data extraction and analysis

Partial data extraction was undertaken by three student research assistants (SS, HA and FB) and completed by two of the authors (EG and AC). For 25 per cent of studies a third team member extracted the data in duplicate. Following data extraction, the studies were sorted by intervention type, categorised into study design type (i.e., model/cost utility analysis or cost-effectiveness analysis), and we considered whether the evidence favoured the intervention or not. Given the heterogeneity of the retained studies meta-analysis was not undertaken. As with most reviews of economic evaluations, the results are presented in the form of a narrative synthesis (9).

### Quality appraisal

We used the Consolidated Health Economic Evaluation Reporting Standards (CHEERS) (10) checklist to assess different aspects of the retained studies’ quality and reporting. The checklist contains 24 items and accompanying recommendations (see Table S3). The overall quality of evidence was assessed using GRADE certainty ratings (11). Final quality was rated high, moderate, low, or very low. The quality assessment was completed by EK. A 25 per cent sample was selected at random and assessed in duplicate by AC. Where results differed, consensus was reached through discussion or the involvement of a third assessor (PM).

## Results

The initial search identified 7386 papers. Duplicates were then removed and after screening by three research assistants (MT, SS, and JP) a further 2462 records were excluded. The random sample of 25% used for checking revealed no conflicts. After the duplicate screening of remaining papers, disagreements (n=26) and records marked as undecided (n=77) were resolved. A total of 59 unique studies met the inclusion criteria (Figure 1). Eight full text papers could not be obtained and reference lists revealed no extra studies. Search 1 identified four grey literature reports, search 2 identified 41 unique studies from the updated Naslund et al (7) search, and search 3 found a further 14 studies of economic models. The non-modelling studies enrolled 5-1514 participants. The USA was the setting for most studies (16), followed by Sweden (10) and the UK (8). For more detailed study characteristics see Table 3.

**Table 2:**
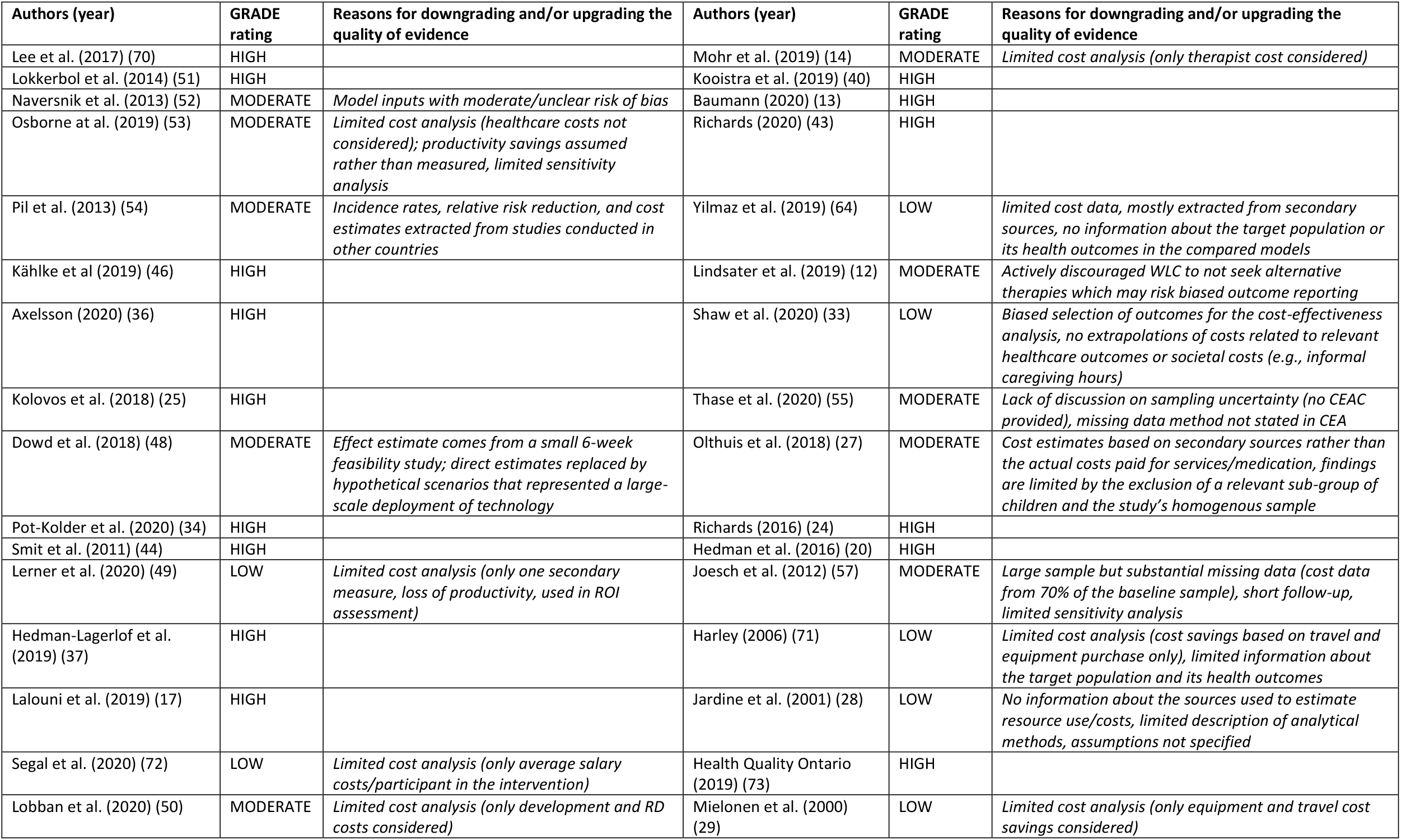
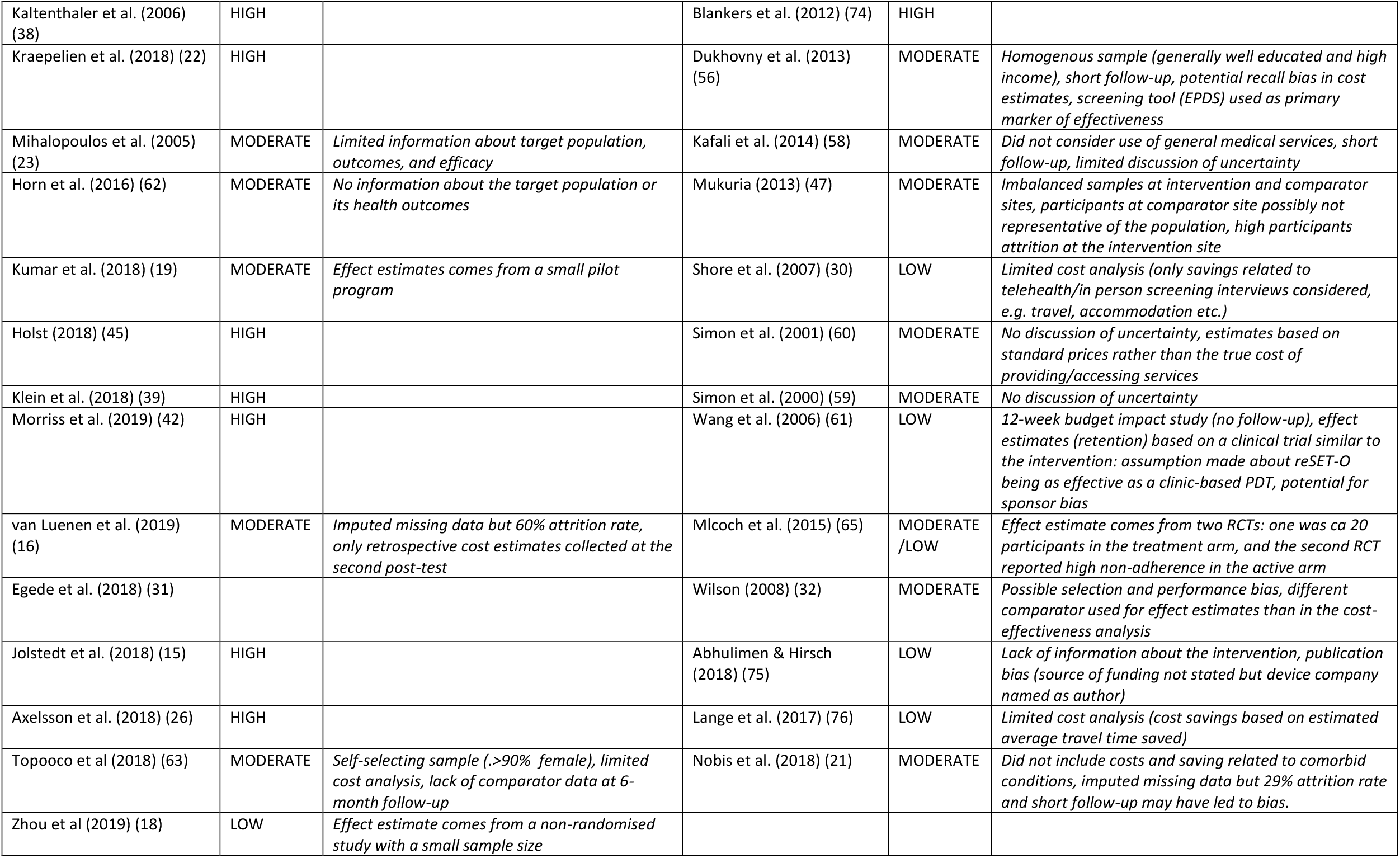
Summary of GRADE certainty ratings.

**Table 3:**
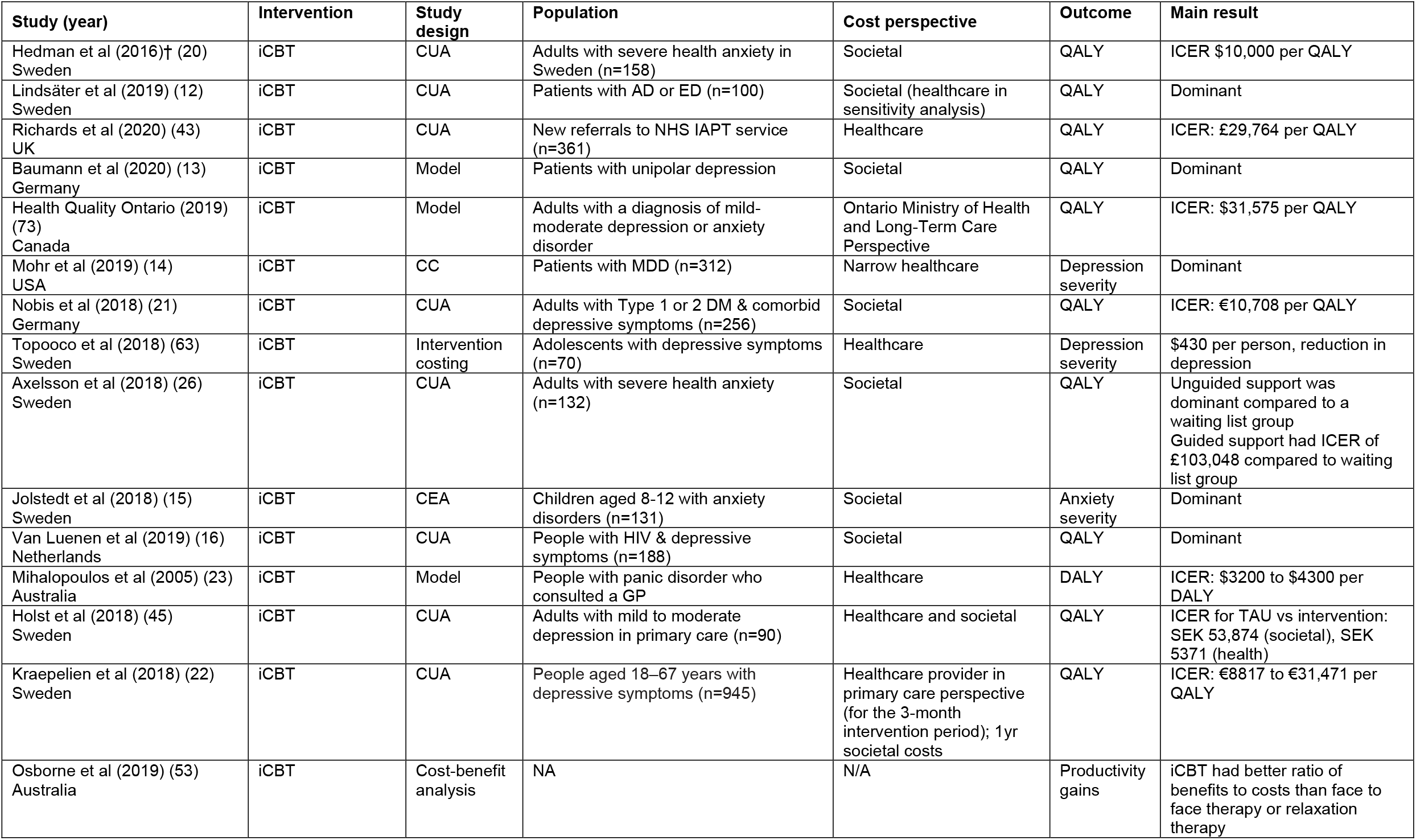
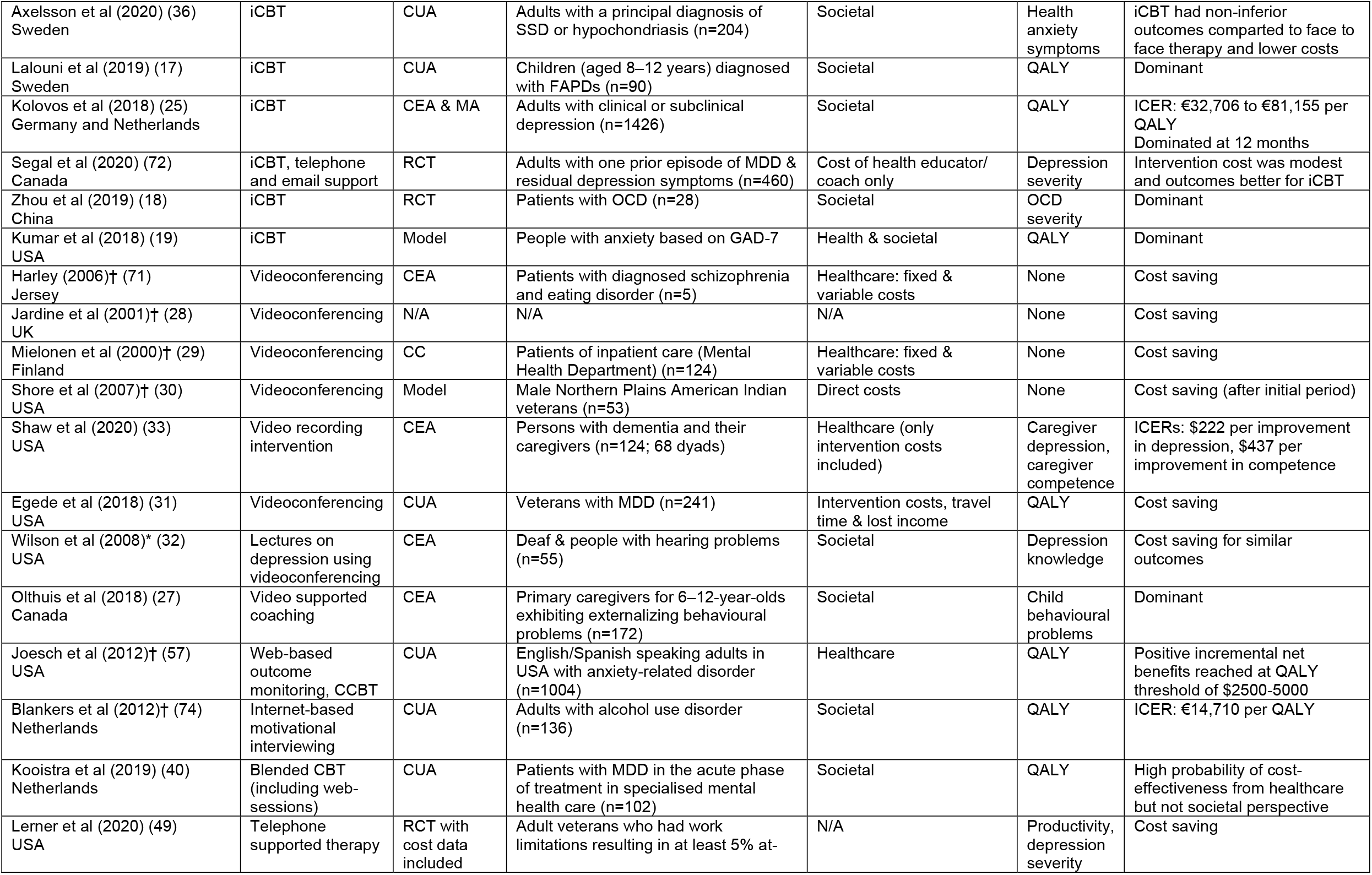
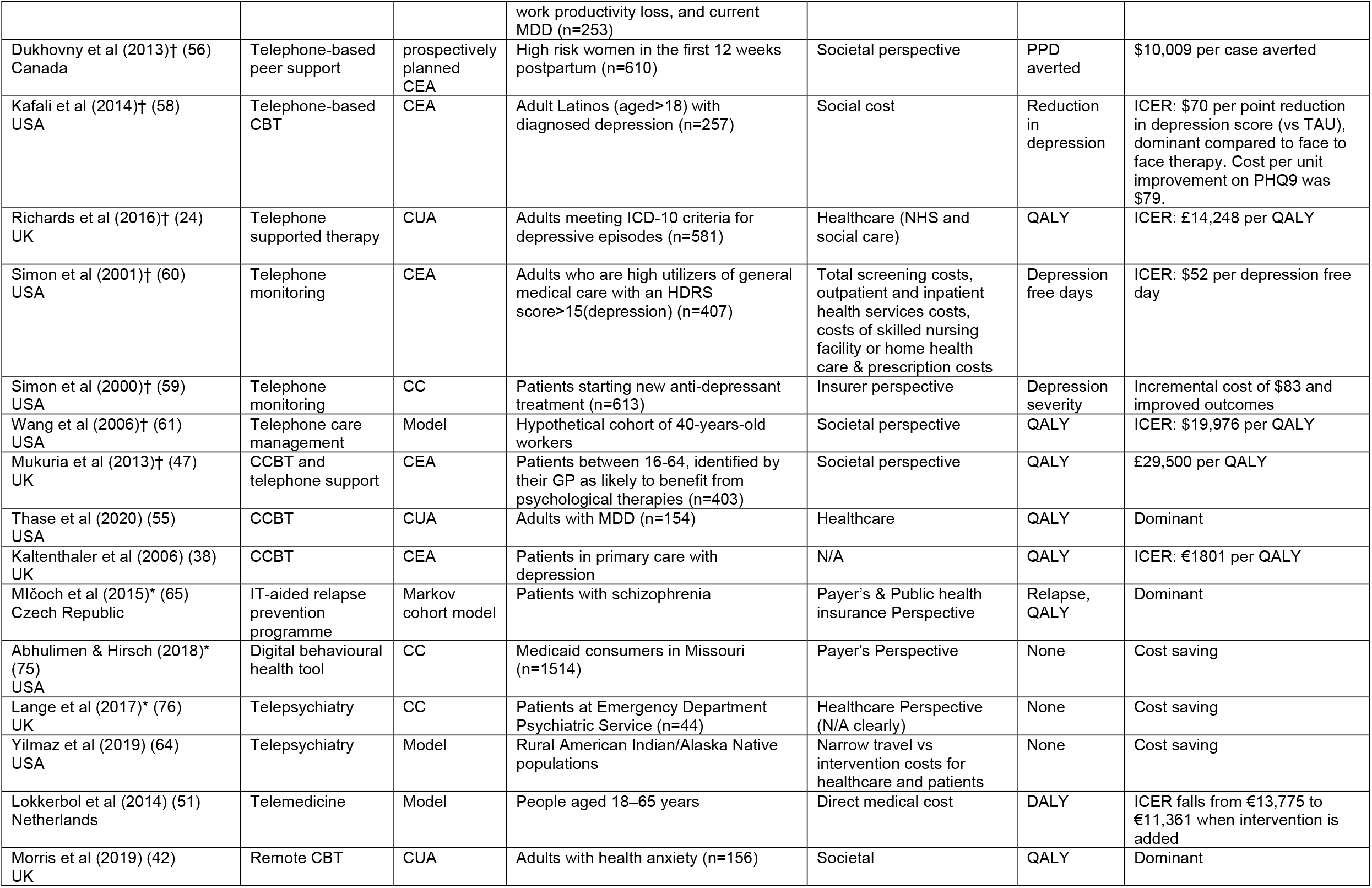
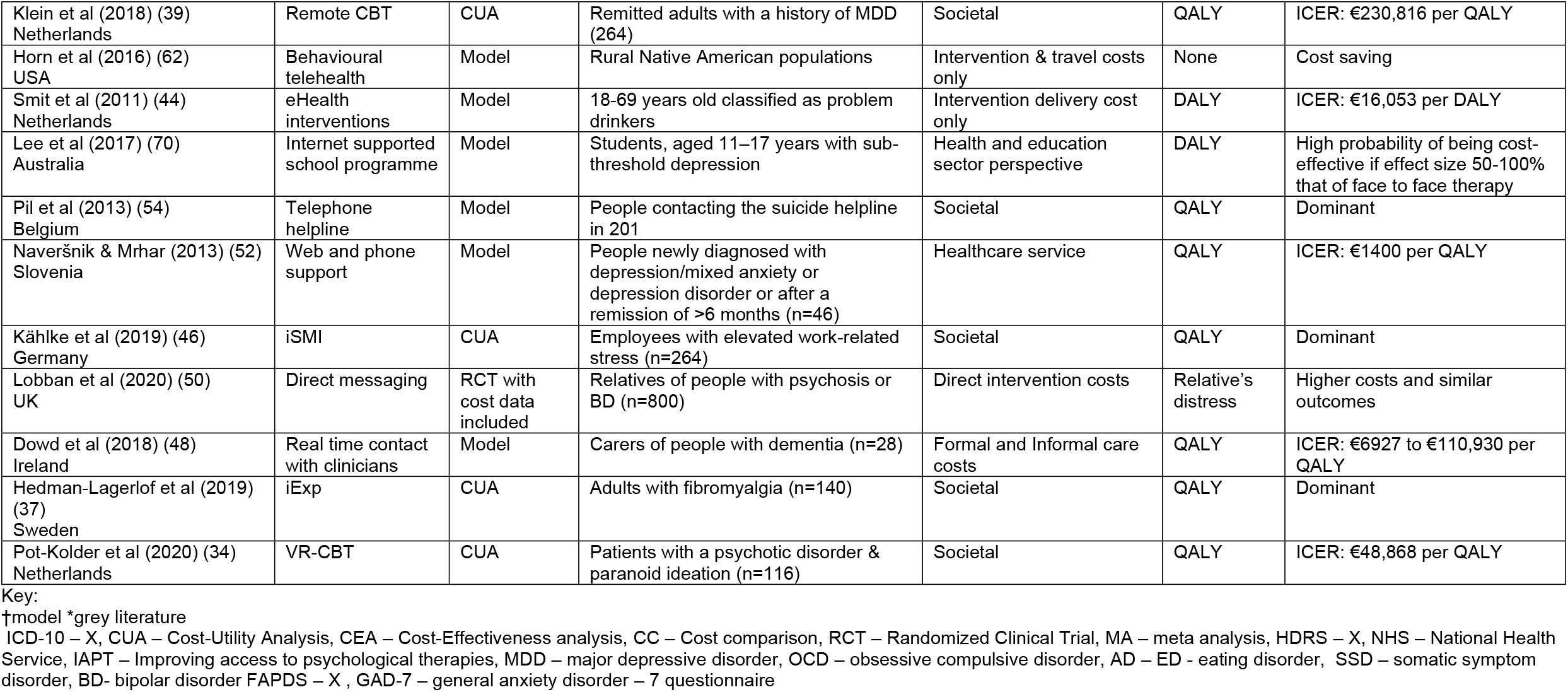
Study characteristics.

The majority (n=40) of included studies found that the telemental health intervention was cost-effective. A further 16 studies indicated that there was some uncertainty around whether the intervention was more cost-effective than the comparator, or that the intervention had potential to be cost-effective but there were some biases in how the study was conducted. Only three studies found that the intervention was less cost-effective than the comparator.

Table 3 shows that the most common interventions were internet-based therapy and videoconferencing. Twenty-one studies evaluated some form of internet-based/delivered therapy (usually cognitive behavioural therapy). QALYs were used in 13 of these studies, DALYs in two studies, productivity losses in one study, and clinically specific measures in the remaining studies. In eight studies the costs for the intervention were lower than the comparator and outcomes were better, indicating a situation of dominance (12–19). In five studies where the intervention had higher costs and produced better outcomes, the ICER was relatively low and indicated cost-effectiveness (20–24). Two studies indicated that therapy was not cost-effective (25, 26).

Videoconferencing and video support to clinicians and patients was evaluated in eight studies. Four of these did not use an outcome measure in the economic analysis and all showed cost savings (27–30). Two other studies reported similar outcomes and lower costs (31, 32). One study, evaluating video coaching for carers of children with behavioural problems, found it to cost saving and outcome improving (27). Another study, found videoconferencing to result in costs per improvement in depression of $222 and per improvement in competence of $437 (33).

Studies that evaluated other remote working interventions usually demonstrated costs savings, dominance (costs savings and better outcomes), or higher costs and better outcomes that justified the extra expense. The study by Pot-Kolder et al (34) found the cost per QALY of virtual reality-based CBT for people with psychosis and paranoid symptoms to be nearly €50,000. However, this was deemed to be cost-effective based on a threshold of €80,000 per QALY for conditions such as this.

The number of studies taking a societal perspective was 27 with 22 taking a healthcare perspective. QALYs were reported in 31 studies and DALYs in four. The reporting quality of each included article was assessed using the 24-item CHEERS checklist: the proportion of studies that met the criteria for each item is shown in Figure 2. 28 studies were of excellent quality, meaning that they addressed satisfactorily at least 90% of applicable checklist items (12, 13, 15–17, 20–22, 24–27, 31, 34–47). 17 studies (14, 19, 23, 48–61) were of very good quality (70-<90%), and further five studies (18, 62–65); had an acceptable quality of reporting (> 60%). The remaining nine studies failed to address or addressed only partially ten or more relevant checklist items. The least reported items were study perspective (item # 6), currency, price date, and conversion (item # 14), the effects of uncertainty (item # 20), and potential conflicts of interest (item # 24). About 40 % of the studies failed to describe fully the characteristics of the base-case population (item # 4), and the sources and methods used to establish clinical effectiveness (item # 11). Similarly, only 80% of the studies reported in sufficient detail the approaches and data sources used to estimate costs (item # 13) and the analytical methods which supported their evaluation (item # 17). Data for subgroup analyses was not available in most cases (item # 21). Finally, approximately half of the studies either did not name its funding source or failed to disclose the funder’s role in the identification, design, conduct, and reporting of the analysis. The detailed performance of each included study for the CHEERS checklist (10) is shown in Table S4 of the Supplement. The overall quality of evidence was assessed using GRADE certainty ratings (11). The assigned ratings shown in Table 2 were based on the quality of the economic evidence.

**Figure 2:**
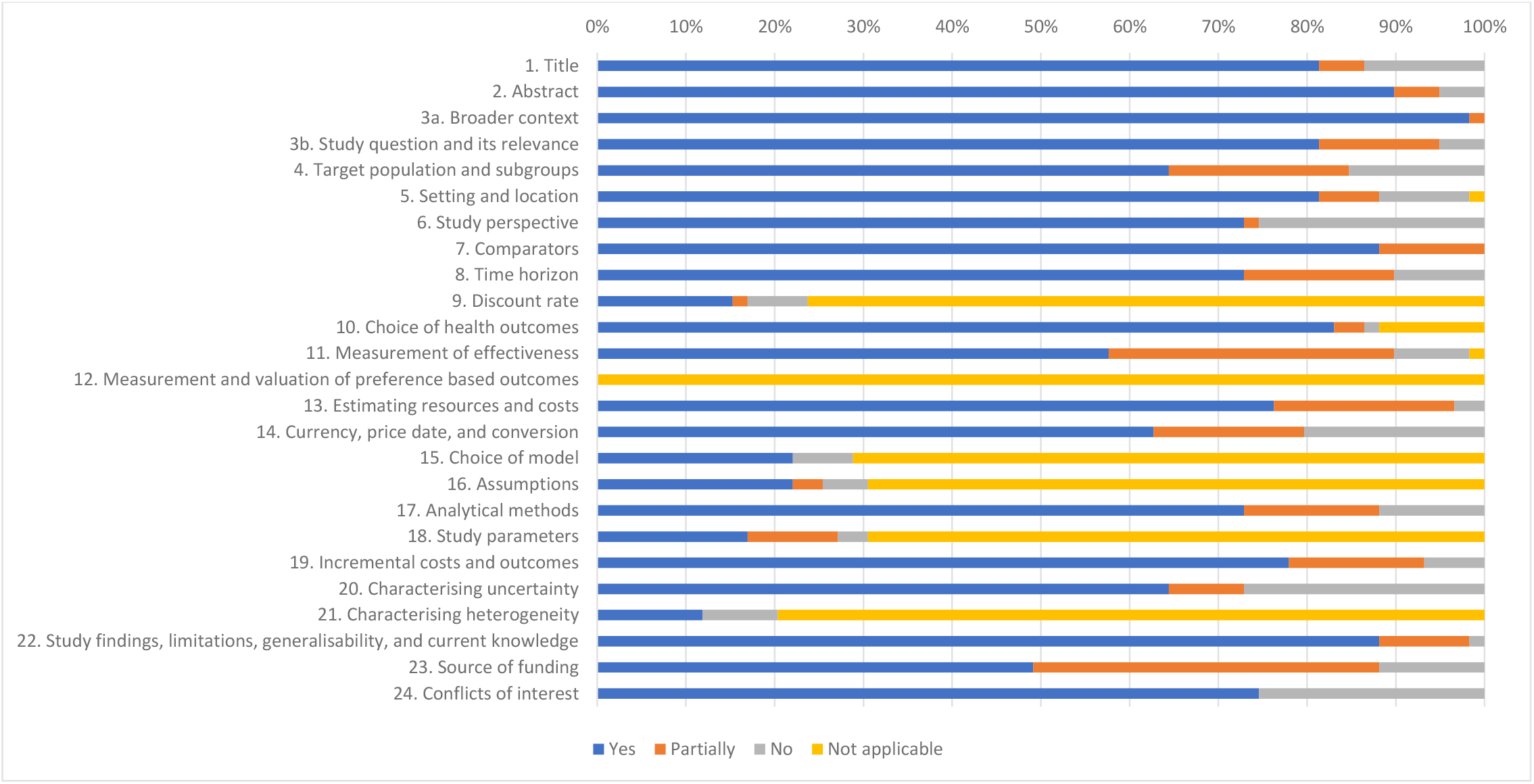
Proportion of included studies that complied with applicable items of the Consolidated Health Economic Evaluation Reporting Standard (CHEERS) checklist.

Overall, there was an over reliance on single-study based estimates. The certainty of evidence was downgraded due to several factors, including imprecision (effect estimates coming from one study with a small sample), risk of bias (narrow perspective, cost omissions related to comorbidity, healthcare utilisation and other relevant aspects, losses to follow-up, short time horizon and limited sensitivity analysis), indirectness (cost estimates based on secondary sources rather than actual resource use, and high percentage of missing data imputed) and potential sponsor bias. The main reasons for downgrading for each study rated as moderate or below are summarised in Table 2.

## Discussion

This systematic review identified 59 studies to build on the findings from Naslund et al (7) regarding the impact of remote technology for mental healthcare interventions in terms of costs and cost-effectiveness. The review by Naslund et al (7) included 26 studies and found good evidence for cost savings associated with telepsychiatry but somewhat limited evidence of cost-effectiveness. We included a broader range of studies and again these suggest that remote working can result in reduced costs but we also identified a reasonable number of full economic evaluations where remote working was either dominant (cost saving and outcome improving) or to result in incremental cost-effectiveness ratios that are below acceptable thresholds. However, as with Naslund et al (7), the studies were markedly heterogeneous with very different designs and perspectives.

The greatest number of studies evaluated some form of internet-delivered therapy (usually cognitive behavioural therapy). These tended to be cost effective as were the various forms of videoconferencing (although many of the latter were limited to cost comparisons).

The perspective taken in the studies was fairly evenly split between a societal perspective (generally meaning that lost work time was included) and a healthcare perspective. This is likely to reflect both the expected benefits of remote working but also the different ways in which healthcare decisions are made in each country. While the latter consideration is important, from an economic point of view we might expect remote working to save patient time and this may include time in work. As such, a societal perspective seems appropriate in this area.

Decision makers often base recommendations on the relationship between costs and QALYs and the studies reviewed here reflect this with 31 using QALYs as the main economic outcome measure. This is interesting especially given some concerns over the use of QALYs in mental health research (66, 67). Most of these concerns though are with the sue of QALYs in studies of interventions for schizophrenia and other severe mental illnesses. As stated above, internet-based cognitive behavioural therapy was most commonly evaluated, and this usually provided for people with depression or anxiety. QALYs do tend to work reasonably well for these conditions.

### Implications

Evaluations of remote working interventions for mental health problems have been criticised for being methodologically limited and this may have held back developments in this area (68). It is from this review that the amount of cost-effectiveness research has increased substantially, and the evidence base is getting stronger. However, methodological issues persist.

The COVID-19 pandemic has clearly led to remote working being given more prominence. While services will to some extent revert back to usual ways of working, some aspects of care delivery that have emerged since early 2020 will most likely remain. In the UK, the charity Mind has identified challenges including the extent to which people have good access to engage with digital approaches, the quality of care delivered in this way not always being of a high standard, and potential breaches of confidentiality (69). As demands on health services continue to increase, it is likely that innovations such as those reviewed here will be needed more and more and it is thus encouraging that on balance they do appear to represent reasonable value for money.

### Limitations

We based our search terms on those used previously by Naslund et al (7). This was appropriate in that we were wanting to update that review but also to include studies from a broader range of sources. However, the strategy may have been too specific and might have missed economic models published prior to 2018. Some terms were excluded that could have been relevant, particularly those relating to the use of social media and platforms such as Zoom or Microsoft Teams.

Another limitation is that some interventions/services only included remote working as a component of a wider package of care or support. It was appropriate to include such studies in order to obtain a broad overview of remote working but identifying the specific impact of remote working is challenging in such studies.

As with other reviews of economic evaluations, included studies were very heterogeneous. This meant that we could only provide a narrative review rather than a more formal synthesis of findings.

## Conclusion

This review has found numerous studies assessing the costs and cost-effectiveness of remote working interventions in mental health. Such approaches appear to be cost-effective although methodological quality of studies needs to be enhanced.

## Data Availability

This is a systematic review and no primary data have been colelcted.

## Data Availability

Data availability is not applicable to this article as no new data were created or analysed in this study.

## Author Contributions

The study was developed by AC and PM. Papers were identified and data extracted by AC, EK, EG, and PM. The manuscript was written by AC, RA, EK, EG and PM.

## Acknowledgements

Joseph Botham assisted with the initial search and data extraction. UCL post-graduate placement students provided invaluable contribution to this study. Initial title and abstract screening was completed by Magdalena Tomoskova, Jiping Mo and Sypros Spyridonids. Data extraction support was completed by Hana Afra, Spyros Spyridonidis and Fatima Bashir.

## Supplemental material

**Table S1:** Naslund et al. (2020) search strategy used in Medline

| Search | Search terms |
| --- | --- |
| #1<br>(Mental disorders) | “serious mental illness” OR “serious and persistent mental illness” OR “severe mental illness” OR “mental illness” OR “mental health” OR “mental disorder” OR “schizophrenia” OR “bipolar disorder” OR “schizoaffective disorder” OR “major depressive disorder” OR “depression” OR “anxiety” OR “affective disorder” OR “psychotic disorders” OR “psychosis” OR “post-traumatic stress disorder” OR “PTSD” OR “stress disorder” OR “antipsychotics” OR “dementia” OR “epilepsy” OR “mental disorders”[MeSH terms] OR “anxiety disorders”[MeSH] OR “stress disorders, traumatic”[MeSH] OR “mood disorders”[MeSH] OR “depressive disorder”[MeSH] OR “schizophrenia and disorders with psychotic features”[MeSH] OR “psychotic disorders”[MeSH] OR “dementia”[MeSH] OR “epilepsy”[MeSH] |
| #2<br>(Substance-use disorders) | “alcohol” OR “substance use” OR “substance use disorder” OR “substance related disorder” OR “alcohol” OR “alcohol use disorder” OR “alcoholism” OR “amphetamine” OR “cocaine” OR “crack cocaine” OR “marijuana” OR “cannabis” OR “opioid” OR “heroin” OR “morphine” OR “street drugs” OR “drug abuse” OR “illicit drug” OR “recreational drugs” OR “substance-related disorders”[MeSH] OR “alcohol-related disorders”[MeSH] OR “amphetamine-related disorders”[MeSH] OR “cocaine- related disorders”[MeSH] OR “marijuana abuse”[MeSH] OR “opioid-related disorders”[MeSH] OR “street drugs”[MeSH] OR “crack cocaine”[MeSH] |
| #3 | #1 OR #2 |
| #4<br>(Digital technology) | “telemetry” OR “telemedicine” OR “telepsychiatry” OR “telehealth” OR “telecare” OR “Tele mental health” OR “connected health” OR “internet” OR “internet health” OR “web browser” OR “website” OR “web-based” OR “social media” OR “Facebook” OR “mobile health” OR “mobile technology” OR “mobile |
|  | phone” OR “cellular phone” OR “cellphone” OR “smartphone” OR “text message” OR “text messaging” OR “wireless technology” OR “remote sensing technology” OR “remote consultation” OR “big data” OR “telemetry”[MeSH] OR “telemedicine”[MeSH] OR “internet”[MeSH] OR “web browser”[MeSH] OR “social media”[MeSH] OR “cellular phone”[MeSH] OR “text messaging”[MeSH] OR “wireless technology”[MeSH] OR “remote sensing technology”[MeSH] |
| #5 (Costs, cost-effectiveness, and economic evaluation) | “cost” OR “cost-effectiveness” OR “cost-benefit” OR “cost-minimisation” OR “cost-utility” OR “eco- nomic” OR “finance” OR “pricing” OR “spending” OR “expenditure” OR “costs and cost analysis”[MeSH] OR “cost-benefit analysis”[MeSH] OR “economics”[MeSH] OR “fees and charges”[MeSH] |
| #6 (Final search) | #3 AND #4 AND #5 |

**Table S2:** Grey literature search strategy

|  |
| --- |
| The grey literature search strategy was simplified from Naslund et al. (2020) search strategy to adapt for grey literature databases. It focused on the following terms:<br>1. tele* OR telehealth OR telepsychiatry OR remote OR technology OR digital<br>2. mental OR “mental health”<br>3. cost* OR economic* OR cost effective<br>4. 1 AND 2 AND 3<br>The term “digital” is not included in the search strategy of Naslund et al. (2020), however it has been added for grey literature searching after it returned relevant results during initial scoping.<br>Where databases had limited search functionality the search terms were further simplified and entered in following priority order:<br>i. telehealth AND mental AND cost<br>ii. tele* AND mental AND cost<br>iii. tele* AND mental AND economic<br>iv. tele* AND mental health AND cost<br>v. tele* AND mental health AND economic<br>vi. telepsychiatry and cost<br>vii. telehealth and cost<br>viii. telehealth and mental<br>ix. remote mental health<br>x. digital mental health<br>xi. technology mental health<br>xii. telepsychiatry<br>xiii. telehealth<br>The following databases and websites were searched on 4-10 November 2020: OpenSIGLE, Healthcare Management Information Consortium, National Technical Information Service, PsycEXTRA, NIHR innovation observatory, Institute of Health Economic, NHSEED, Substance Abuse & Mental Services Administration Evidence-Based Practices Resource Center, British Library, CADTH, Current Awareness Service for Health COPAC, EThOS, NDLTD, Nuffield Trust, OpenGrey, ISPOR conference, The Health Foundation. |

**Table S3:** Consolidated Health Economic Evaluation Reporting Standards (CHEERS) checklist

| Section/Item | No | Recommendation |
| --- | --- | --- |
| **Title and abstract** |  |  |
| Title | 1 | Identify the study as an economic evaluation or use more specific terms such as “cost-effectiveness analysis”, and describe the interventions compared. |
| Abstract | 2 | Provide a structured summary of objectives, perspective, setting, methods (including study design and inputs), results (including base case and uncertainty analyses), and conclusions. |
| **Introduction** |  |  |
| Background and objectives | 3a | Provide an explicit statement of the broader context for the study. |
|  | 3b | Present the study question and its relevance for health policy or practice decisions. |
| Methods |  |  |
| Target population and subgroups | 4 | Describe characteristics of the base case population and subgroups analysed, including why they were chosen. |
| Setting and location | 5 | State relevant aspects of the system(s) in which the decision(s) need(s) to be made. |
| Study perspective | 6 | Describe the perspective of the study and relate this to the costs being evaluated. |
| Comparators | 7 | Describe the interventions or strategies being compared and state why they were chosen. |
| Time horizon | 8 | State the time horizon(s) over which costs and consequences are being evaluated and say why appropriate. |
| Discount rate | 9 | Report the choice of discount rate(s) used for costs and outcomes and say why appropriate. |
| Choice of health outcomes | 10 | Describe what outcomes were used as the measure(s) of benefit in the evaluation and their relevance for the type of analysis performed. |
| Measurement of effectiveness | 11a | Single study-based estimates: Describe fully the design features of the single effectiveness study and why the single study was a sufficient source of clinical effectiveness data. |
|  | 11b | Synthesis-based estimates: Describe fully the methods used for identification of included studies and synthesis of clinical effectiveness data. |
| Measurement and valuation of preference based outcomes | 12 | If applicable, describe the population and methods used to elicit preferences for outcomes. |
| Estimating resources and costs | 13a | Single study-based economic evaluation: Describe approaches used to estimate resource use associated with the alternative interventions. Describe primary or secondary research methods for valuing each resource item in terms of its unit cost. Describe any adjustments made to approximate to opportunity costs. |
|  | 13b | Model-based economic evaluation: Describe approaches and data sources used to estimate resource use associated with model health states. Describe primary or secondary research methods for valuing each resource item in terms of its unit cost. Describe any adjustments made to approximate to opportunity costs. |
| Currency, price date, and conversion | 14 | Report the dates of the estimated resource quantities and unit costs. Describe methods for adjusting estimated unit costs to the year of reported costs if necessary. Describe methods for converting costs into a common currency base and the exchange rate. |
| Choice of model | 15 | Describe and give reasons for the specific type of decision-analytical model used. Providing a figure to show model structure is strongly recommended. |
| Assumptions | 16 | Describe all structural or other assumptions underpinning the decision-analytical model. |
| Analytical methods | 17 | Describe all analytical methods supporting the evaluation. This could include methods for dealing with skewed, missing, or censored data; extrapolation methods; methods for pooling data; approaches to validate or make adjustments (such as half cycle corrections) to a model; and methods for handling population heterogeneity and uncertainty. |
| **Results** |  |  |
| Study parameters | 18 | Report the values, ranges, references, and, if used, probability distributions for all parameters. Report reasons or sources for distributions used to represent uncertainty where appropriate. Providing a table to show the input values is strongly recommended. |
| Incremental costs and outcomes | 19 | For each intervention, report mean values for the main categories of estimated costs and outcomes of interest, as well as mean differences between the comparator groups. If applicable, report incremental cost-effectiveness ratios. |
| Characterising uncertainty | 20a | Single study-based economic evaluation: Describe the effects of sampling uncertainty for the estimated incremental cost and incremental effectiveness parameters, together with the impact of methodological assumptions (such as discount rate, study perspective). |
|  | 20b | Model-based economic evaluation: Describe the effects on the results of uncertainty for all input parameters, and uncertainty related to the structure of the model and assumptions. |
| Characterising heterogeneity | 21 | If applicable, report differences in costs, outcomes, or cost-effectiveness that can be explained by variations between subgroups of patients with different baseline characteristics or |
|  |  | other observed variability in effects that are not reducible by more information. |
| **Discussion** |  |  |
| Study findings, limitations, generalisability, and current knowledge | 22 | Summarise key study findings and describe how they support the conclusions reached. Discuss limitations and the generalisability of the findings and how the findings fit with current knowledge. |
| **Other** |  |  |
| Source of funding | 23 | Describe how the study was funded and the role of the funder in the identification, design, conduct, and reporting of the analysis. Describe other non-monetary sources of support. |
| Conflicts of interest | 24 | Describe any potential for conflict of interest of study contributors in accordance with journal policy. In the absence of a journal policy, we recommend authors comply with International Committee of Medical Journal Editors recommendations. |

Source: Husereau D, Drummond M, Petrou S, Carswell C, Moher D, Greenberg D et al. (2013) Consolidated Health Economic Evaluation Reporting Standards (CHEERS) statement. BMJ 346 :f1049 doi:10.1136/bmj.f1049.

**Table S4.**
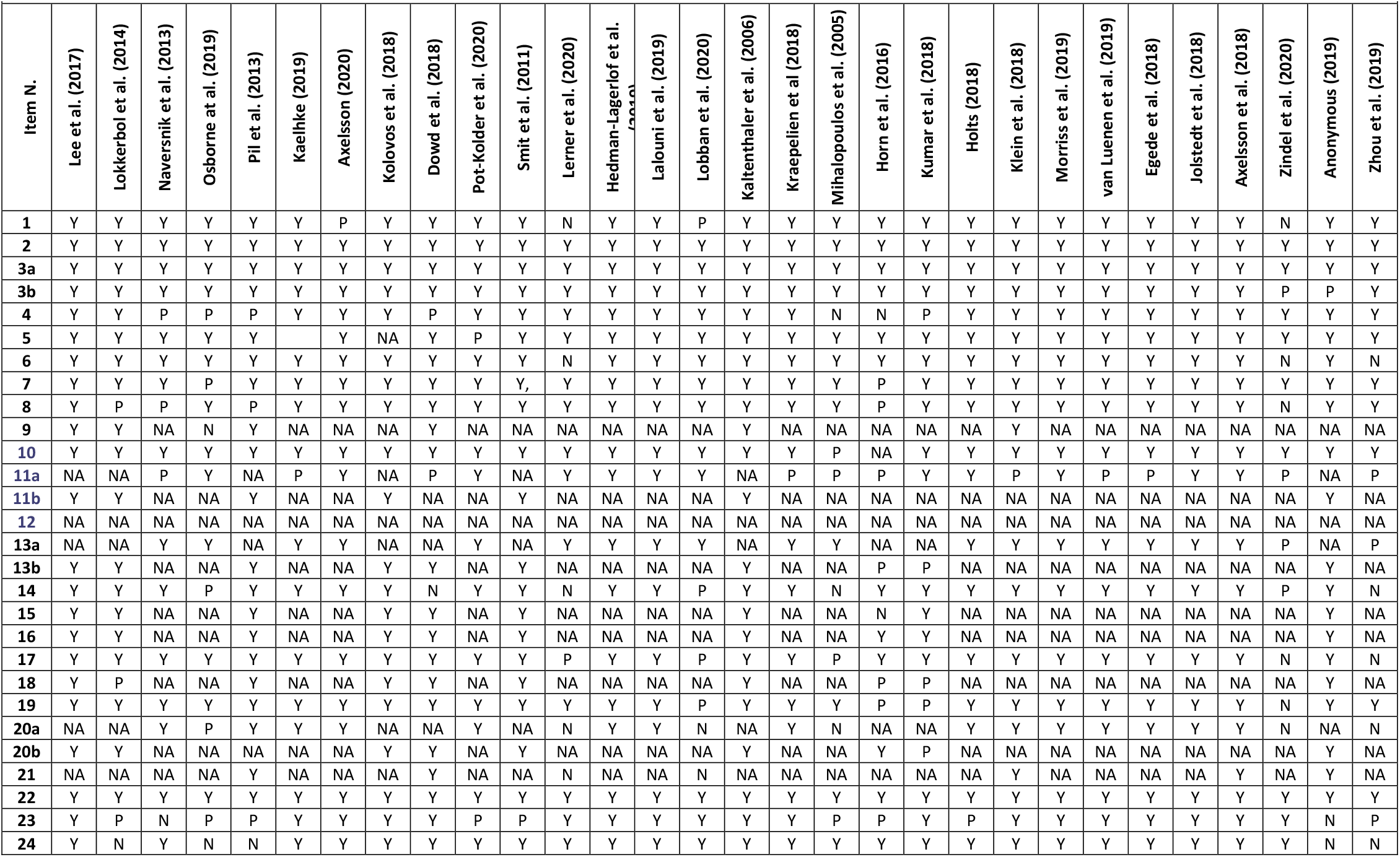
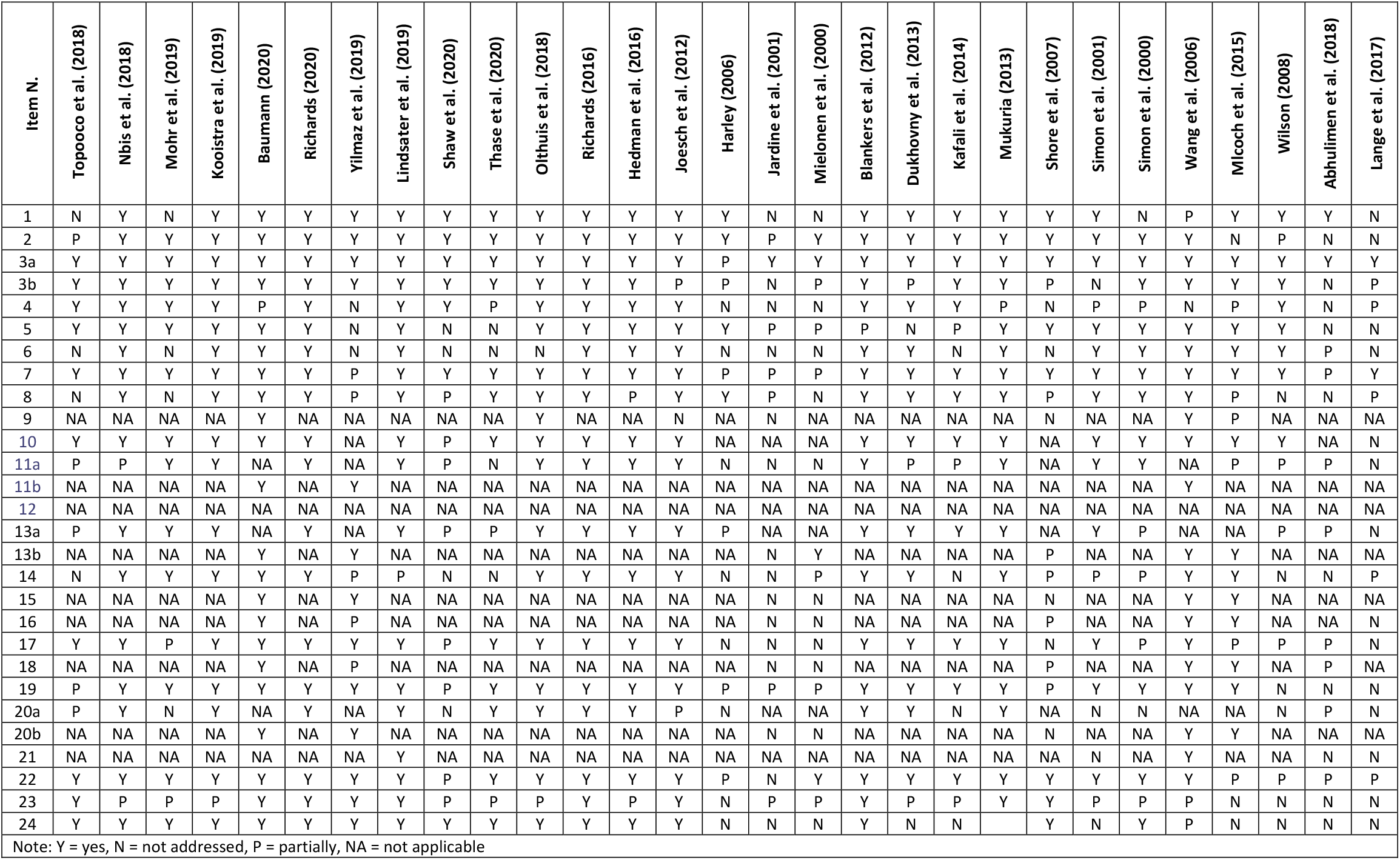
CHEERS checklist: Results

## Footnotes

1 The following databases used in Naslund et al. (2020) were not included in this review due to organisational access and the number of duplicates already identified: Web of Science, Health Economic Evaluations Database, Cost-Effectiveness Analysis Registry, Research Papers in Economics (RcPEc), and European Network of Health Economic Evaluation Database (EURONHEED).

2 In CINAHL we were unable to apply MeSH terms, and in EconLit the simplified grey literature strategy was used (see Table S2).

